# Epidemiological and Genomic Analysis of SARS-CoV-2 in Ten Patients from a Mid-sized City outside of Hubei, China in the Early Phase of the COVID-19 Outbreak

**DOI:** 10.1101/2020.04.16.20058560

**Authors:** Jinkun Chen, Evann E. Hilt, Huan Wu, Zhuojing Jiang, QinChao Zhang, JiLing Wang, Yifang Wang, Fan Li, Ziqin Li, Jialiang Tang, Shangxin Yang

## Abstract

A novel coronavirus known as severe acute respiratory syndrome coronavirus 2 (SARS-CoV-2) is the cause of the ongoing COVID-19 pandemic. In this study, we performed a comprehensive epidemiological and genomic analysis of SARS-CoV-2 genomes from ten patients in Shaoxing (Zhejiang Province), a mid-sized city outside of the epicenter Hubei province, China, during the early stage of the outbreak (late January to early February, 2020). We obtained viral genomes with > 99% coverage and a mean depth of 296X demonstrating that viral genomic analysis is feasible via metagenomics sequencing directly on nasopharyngeal samples with SARS-CoV-2 Real-time PCR C_t_ values less than 28. We found that a cluster of 4 patients with travel history to Hubei shared the exact same virus with patients from Wuhan, Taiwan, Belgium and Australia, highlighting how quickly this virus spread to the globe. The virus from another cluster of two family members living together without travel history but with a sick contact of a confirmed case from another city outside of Hubei accumulated significantly more mutations (9 SNPs vs average 4 SNPs), suggesting a complex and dynamic nature of this outbreak. Our findings add to the growing knowledge of the epidemiological and genomic characteristics of SARS-CoV-2 and offers a glimpse into the early phase of this viral infection outside of Hubei, China.

## INTRODUCTION

Coronaviruses (CoVs) are a large family of single-stranded RNA viruses that can be isolated from a variety of animals including camels, rats, birds and bats (Cascella et al., 2020). These coronaviruses can cause a range of disease states in animals including respiratory, enteric, hepatic and neurological disease (Weiss and Leibowitz, 2011). Before late 2019, there were six known CoVs capable of infecting humans (Hu-CoVs). The first four Hu-CoVs that cause mild disease are HKU1, NL63, OC43 and 229E and are known to circulate in the human population (Corman et al., 2018). The other two Hu-CoVs, known as severe acute respiratory syndrome-CoV (SARS-CoV) and middle east respiratory syndrome-CoV (MERS-CoV), caused two previous epidemics in 2003 (Rota et al., 2003) and 2012 (Zaki et al., 2012) respectively. Both SARS-CoV and MERS-CoV were the results of recent spillover events from animals. These two epidemics highlighted how easy it is for spillover events in CoVs to occur and cause outbreaks in humans.

In December 2019, another spillover event occurred and a seventh Hu-CoV appeared known as severe acute respiratory syndrome coronavirus 2 (SARS-CoV-2), previously named 2019-nCoV (Wu et al., 2020). SARS-CoV-2 has been spreading rapidly across the world since it was first reported in Wuhan, Hubei province, China (Wang et al., 2020a; Wu et al., 2020). The advances and accessibility of sequencing technologies have allowed researchers all over the world to quickly sequence the genome of SARS-CoV-2 (Shu and McCauley, 2017; Zhou et al., 2020). Zhou *et al*. 2020 showed that SARS-CoV-2 shared 79.6% sequence identity to SARS-CoV and 96% sequence identity to a bat CoV further supporting the theory of another spillover event (Zhou et al., 2020).

Genomic analysis of SARS-CoV-2 genomes suggested that there were two major genotypes in the early phase of the outbreak, known as L type and S type, based on almost complete linkage between two SNPs (Tang et al., 2020). Tang *et al*. 2020 proposed that the L type may be more aggressive in replication rates and spreads more quickly (Tang et al., 2020). A recent reclassification of SARS-CoV-2 was proposed, and most viruses of the S type and L type described in the Tang’s study are now classified as Lineage A and B, respectively (Rambaut et al., 2020). Here we present a comprehensive epidemiological and genomic analysis of SARS-CoV-2 genomes from 10 patients in Shaoxing (Zhejiang Province), a mid-sized city about 500 miles away from Wuhan at the early stages of the outbreak.

## MATERIALS AND METHODS

### Study design and Ethics

Ten remnant nasopharyngeal swab samples collected between 1/27/2020 and 2/7/2020, and tested positive by a SARS-CoV-2 real-time PCR assay with cycle threshold (C_t_) values of less than 28, were included in this study. The samples were de-identified except the associated epidemiological data were retained. Since the patient identification was removed and the samples used in this study were remnant and otherwise would be discarded, the Shaoxing Center for Disease Control and Prevention had determined that the institutional review boards (IRB) approval was waived for this project, and the informed consent form was not required.

### SARS-CoV-2 PCR & RNA Sequencing

Total nucleic acid was extracted from the nasopharyngeal swabs using the Total Nucleic Acid Extraction Kit (IngeniGen XMK Biotechnologies, Inc. Zhejiang, China). Real-time PCR was performed by using the IngeniGen XMKbio 2019-nCoV (SARS-CoV-2) RNA Detection kit, which targets the highly specific sequences in the *ORF1ab* and *N* genes of the virus, on the ABI 7500 system (ThermoFisher Scientific, Inc. MA, USA). The RNA libraries were constructed using the Ingenigen XMKbio RNA-seq Library Prep Kit (IngeniGen XMK Biotechnologies, Inc. Zhejiang, China). Briefly, DNase was used to remove residual human DNA and the RNA was fragmented, followed by double-strand cDNA synthesis, end-repair, dA-tailing and adapter ligation. Sequencing was performed by using the 2X75bp protocol on the Nextseq 550 system (Illumina, Inc. CA, USA).

### Data Analysis

Quality control and trimming of paired-end reads was performed using custom Python scripts as follows: 1) trim 3’ adapters; 2) trim reads at ambiguous bases; 3) filter reads shorter than 40bp; 4) filter reads with average quality score < 20. Host-derived reads were removed by alignment against the GRCh38.p13 genome reference using bowtie2 (v2.3.4.3) with default parameters. The retained reads were then mapped to 163 published SARS-CoV-2 reference genomes obtained from GISAID (https://www.gisaid.org/CoV2020/, accessed March 2, 2020) by bowtie2 (v2.3.4.3) with default parameters. snippy (v4.5.0) was used for variant calling and core SNP alignment against the Wuhan-Hu-1 reference, FastTree (v2.1.3) was used for tree construction using default parameters, and Figtree (v1.4.4) was used to visualize the resulting phylogenetic tree. Additional statistical analyses and visualizations were performed using custom Python scripts with the pandas (v0.25.0) and matplotlib (v3.1.1) modules. The L/S type was determined according to methods previously described (Tang et al., 2020). Briefly, C at position 8782 and T at position 28144 was determined to be A type, and T at position 8782 and C at position 28144 was determined to be B type. The A/B lineage was determined by using pipeline pangolin (https://github.com/hCoV-2019/pangolin) as described previously (Rambaut et al., 2020).

## RESULTS

### Epidemiology of Shaoxing Patients

All ten patients presented with symptoms (fever and cough) consistent with COVID-19 in late January and early February of 2020. The majority of patients were male (60%) and the average age was 44 (**Table 1**). The patients can be categorized into two epidemiologic groups with either a travel history to the Hubei province or sick contact with a confirmed case (**Table 1**). There was one case where we were unable to obtain a travel or exposure history (Shaoxing-8).

**Table 1.** Epidemiological History of the 10 Shaoxing Patients.

| ID | Age Range | History of Travel or Sick Contact | Date of Symptom Onset | Date of Sample Collection |
| --- | --- | --- | --- | --- |
| Shaoxing-01 | 30-39 | Family members traveled together to Hubei province (1/15 – 1/24) | 1/24/20 | 1/27/20 |
| Shaoxing-02 | 70-79 |  | 1/29/20 | 1/30/20 |
| Shaoxing-03 | 60-66 |  | 1/28/20 | 1/30/20 |
| Shaoxing-04 | 50-59 |  | 1/29/20 | 1/31/20 |
| Shaoxing-05 | 50-59 | Traveled to Hubei (1/16 – 1/23) | 1/29/20 | 1/31/20 |
| Shaoxing-06 | 30-39 | Resident of Wuhan;<br>Traveled to Shaoxing on 1/17 | 1/29/20 | 1/31/20 |
| Shaoxing-07 | <10 | Traveled to Hubei (1/11 – 1/24);<br>Two family members were confirmed cases | 1/30/20 | 1/30/20 |
| Shaoxing-08 | 50-59 | Unknown | 1/31/20 | 2/7/20 |
| Shaoxing-09 | 30-39 | Family members living together no travel history;<br>Contact with a confirmed case from Ningbo, Zhejiang on 1/27 | 2/2/20 | 2/5/20 |
| Shaoxing-10 | 30-39 |  | 2/5/20 | 2/6/20 |

There are two apparent clusters in these ten patients. The first cluster involves four patients who are relatives and traveled together to Hubei province for a wedding in late January. The first patient in this cluster had symptom onset on their last day in Hubei province while the other three patients had symptom onset 4-5 days after coming back to Shaoxing (**Table 1**). The second cluster involves two patients who are family members that live together and did not travel to Hubei province. One of the family members (Shaoxing-09) had a sick contact with a confirmed case who visited her but lived in Ningbo, a more populated city in Zhejiang province (**Table 1**).

### Metagenomic Sequencing

The patients were confirmed to have SARS-CoV-2 infection by a commercial Real-time PCR assay. The average C_t_ values for the 10 patient samples were 23.17 for *ORF1ab* and 24.54 for *N* (**Table 2**). Metagenomic sequencing was performed to recover the full viral genome. The total number of sequence reads per samples ranged from 10.4 million to 27.5 million with an average of 17.1 million. A small percentage of these reads mapped to SARS-CoV-2 RNA (**Table 2**). The range of sequence reads that mapped to SARS-CoV-2 RNA was 2,413 to 163,158 with an average of 49,066. We observed a clear negative correlation between the C_t_ values of each gene (*ORF1ab* and *N*) and the log value of SARS-CoV-2 RNA reads (Figure 1). However, the linearity is not significant (R^2^=0.6628, 0.5595 for *ORF1ab* and *N*, respectively), indicating that the number of RNA reads measured by metagenomics sequencing are only semi-quantitative and cannot be interpreted directly as viral loads.

**Table 2.** Summary of Sequencing Results of 10 Shaoxing Patient Samples.

| ID | Ct Value (ORF1ab) | Ct Value (N) | Total Reads (PE 75) | 2019-nCoV RNA (Raw Reads) | 2019-nCoV RNA (Log Value) | Genome Coverage (%) | Mean Depth (X) |
| --- | --- | --- | --- | --- | --- | --- | --- |
| Shaoxing-01 | 21.57 | 23.62 | 17,158,277 | 40,057 | 4.60 | 99.4 | 219 |
| Shaoxing-02 | 18.86 | 20.93 | 13,602,710 | 149,682 | 5.18 | 99.9 | 929 |
| Shaoxing-03 | 20.09 | 22.25 | 24,769,343 | 163,158 | 5.21 | 99.8 | 1024 |
| Shaoxing-04 | 24.02 | 24.68 | 21,509,477 | 15,424 | 4.19 | 100.0 | 81 |
| Shaoxing-05 | 21.81 | 23.81 | 14,043,326 | 99,521 | 5.00 | 99.9 | 591 |
| Shaoxing-06 | 25.88 | 27.08 | 18,909,299 | 3,535 | 3.55 | 99.9 | 18 |
| Shaoxing-07 | 23.11 | 23.87 | 10,480,051 | 5,063 | 3.70 | 99.8 | 26 |
| Shaoxing-08 | 23.34 | 24.53 | 11,506,909 | 2,413 | 3.38 | 99.9 | 12 |
| Shaoxing-09 | 26.81 | 27.85 | 27,517,291 | 8,897 | 3.95 | 99.9 | 47 |
| Shaoxing-10 | 26.24 | 26.8 | 11,071,595 | 2,911 | 3.46 | 99.7 | 15 |
| Min | 18.86 | 20.93 | 10,480,051 | 2,413 | 3.38 | 99.4 | 12 |
| Max | 26.81 | 27.85 | 27,517,291 | 163,158 | 5.21 | 100.0 | 1024 |
| Mean | 23.17 | 24.54 | 17,056,828 | 49,066 | 4.22 | 99.8 | 296 |

**Figure 1.**
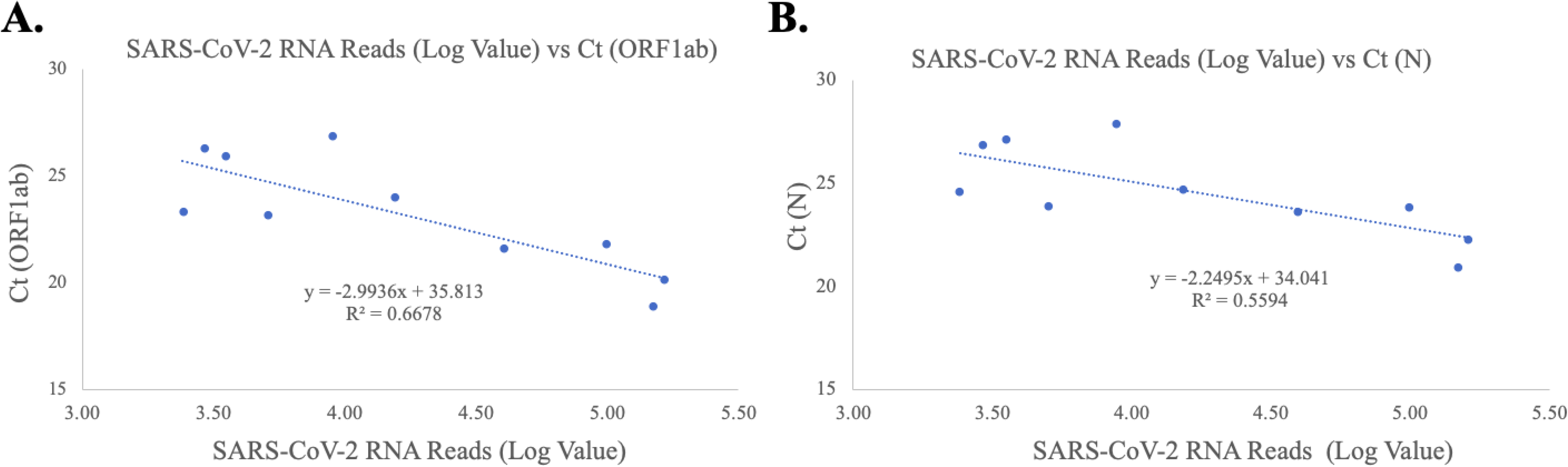
Correlation of C_t_ Values and SARS-CoV-2 RNA Reads. **(a) *ORF1ab* Gene Target**. The X-axis plots the log value of the SARS-CoV-2 RNA reads while the Y-axis plots the C_t_ values for the *ORF1ab* gene for the ten Shaoxing patients. **(b) *N* Gene Target**. The X-axis plots the log value of the SARS-CoV-2 RNA reads while the Y-axis plots the C_t_ values for the *N* gene for the ten Shaoxing patients.

With a large variation in the SARS-CoV-2 RNA mapped reads, we were still able to obtain excellent coverage and depth when each genome was mapped to the first SARS-CoV-2 genome, Wuhan-Hu-1 (Wu et al., 2020) (**Figure 2A**). The coverage for all genomes was above 99% and the mean depth for the genomes ranged from 12X to 1024X (**Table 2, Figure 2B**). Genomes sequenced to a relatively low mean depth (12X to 47X) were still able to be genotyped successfully (see Results below) but our results suggest that SARS-CoV-2 read counts of at least 15,000 yield sufficiently high depth to characterize even low prevalence or rare mutations.

**Figure 2.**
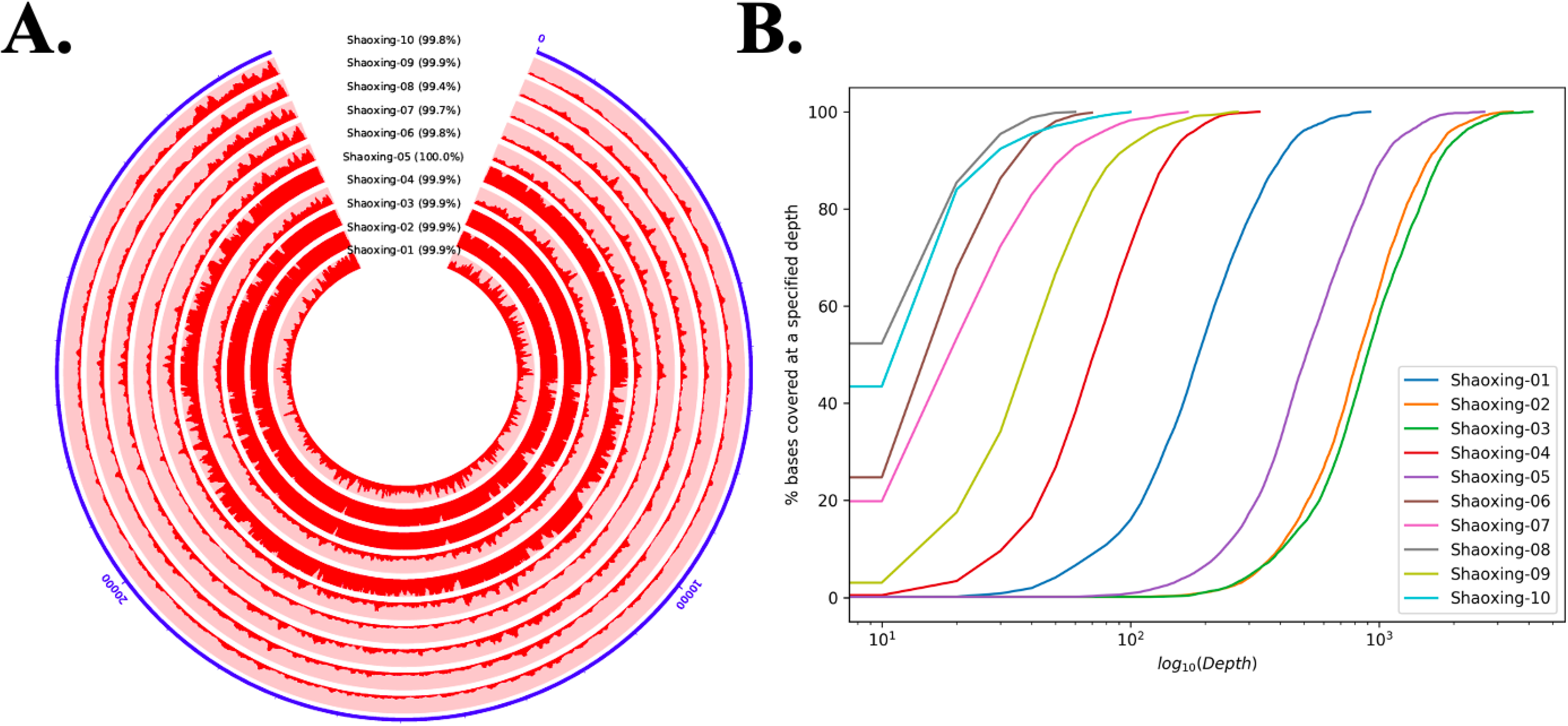
Coverage and Depth. **(a) Coverage and Depth Map**. The coverage and depth at each base are depicted by the dark red shading along the circle. **(b) Depth Ratio**. The X-axis plots the log value of the depth for each genome while the Y-axis plots the cumulative percentage of bases covered to the specified depth.

### Estimation of Mutation Rate

To determine the single nucleotide polymorphisms (SNPs) of SARS-CoV-2 in these 10 patients, we mapped each genome to the original Wuhan-Hu-1 reference which was collected on December 31, 2019 (Wu et al., 2020). The genomes contained a fairly moderate number of SNPs (mean of 4 SNPs, range 1-9) (**Table 3**), consistent with previous reports of relatively low mutation rates (Wang et al., 2020b). The genomes with the largest number of SNPs came from individuals who had contact with a confirmed case from Ningbo, Zhejiang and no travel history to the Hubei province (**Table 3**, **Shaoxing-9 and 10**).

**Table 3.** Summary of Genomic Descriptions for the Shaoxing SARS-CoV-2 Genomes.

| ID | Lineage | Type | No. of SNP<sup>a</sup> | No. of Days<sup>b</sup> | Mutation Rate<br>(#SNP/day) | Mutation Rate<br>(#SNP/day/nt) | Mutation Rate<br>(#SNP/yr/nt) |
| --- | --- | --- | --- | --- | --- | --- | --- |
| Shaoxing-01 | A | S | 2 | 27 | 0.07 | 2.48E-06 | 9.04E-04 |
| Shaoxing-02 | A | S | 2 | 30 | 0.07 | 2.23E-06 | 8.14E-04 |
| Shaoxing-03 | A | S | 2 | 30 | 0.07 | 2.23E-06 | 8.14E-04 |
| Shaoxing-04 | A | S | 2 | 31 | 0.06 | 2.16E-06 | 7.87E-04 |
| Shaoxing-05 | B | L | 2 | 31 | 0.06 | 2.16E-06 | 7.87E-04 |
| Shaoxing-06 | A | S | 3 | 31 | 0.10 | 3.24E-06 | 1.18E-03 |
| Shaoxing-07 | B | L | 5 | 30 | 0.17 | 5.57E-06 | 2.03E-03 |
| Shaoxing-08 | B | L | 1 | 38 | 0.03 | 8.80E-07 | 3.21E-04 |
| Shaoxing-09 | A | S | 9 | 36 | 0.25 | 8.36E-06 | 3.05E-03 |
| Shaoxing-10 | A | S | 9 | 37 | 0.24 | 8.13E-06 | 2.97E-03 |
| Min |  |  | 1 | 27 | 0.03 | 8.80E-07 | 3.21E-04 |
| Max |  |  | 9 | 38 | 0.25 | 8.36E-06 | 3.05E-03 |
| Mean |  |  | 4 | 32 | 0.11 | 3.74E-06 | 1.37E-03 |

<sup>a</sup> SNP calculated by mapping each genome to the genome of Wuhan-Hu-1 (Wu et al., 2020)
<sup>b</sup> Number of days between the date that the sample was collected and the date the Wuhan-Hu-1 genome was published (12/31/2019)

Using the SNP analysis, we calculated the various mutation rates using the number of days between the date that the sample was collected and the date the Wuhan-Hu-1 sample was collected. The mutation rate (SNP per day) ranged from 0.03 to 0.25 (**Table 3**). We used this mutation rate to calculate the nucleotide substitution per site per day and the nucleotide substitution per site per year. We saw an average mutation rate of 3.74×10^-6^ nucleotide substitution per site per day and an average mutation rate of 1.37×10^-3^ nucleotide substitution per site per year (**Table 3**).

We investigated each SNP to determine if there were any non-synonymous mutations in genes important to the virus lifecycle (**Table 4**). No non-synonymous mutations were found in the S gene, which encodes the spike protein that’s critical for viral binding to human receptor ACE2 (Zhou et al., 2020). Notably in the cluster of the two family members (Shaoxing-9 and - 10), the two viruses are closely related but not identical. Shaoxing-9 was infected first and then transmitted to Shaoxing-10, whose virus gained a non-synonymous mutation C9962T in the ORF1ab gene (**Table 4**). This could be explained by the sequential transmission, however, we could not rule out a possibility of intra-host viral heterogeneity in the two patients.

**Table 4.** Summary of SNPs in the Ten SARS-CoV-2 Genomes.

| SNP# | Position | Gene | Reference nt | Shaoxing-01 | Shaoxing-02 | Shaoxing-03 | Shaoxing-04 | Shaoxing-05 | Shaoxing-06 | Shaoxing-07 | Shaoxing-08 | Shaoxing-09 | Shaoxing-10 |
| --- | --- | --- | --- | --- | --- | --- | --- | --- | --- | --- | --- | --- | --- |
| 1 | 207 | non-coding | C |  |  |  |  |  |  |  |  | T (non-coding) |  |
| 2 | 889 | orf1ab | T |  |  |  |  |  |  | C (A) |  |  |  |
| 3 | 946 | orf1ab | T |  |  |  |  |  |  |  |  | C (G) | C (G) |
| 4 | 5099 | orf1ab | T |  |  |  |  |  |  |  |  | A (S->T) | A (S->T) |
| 5 | 7420 | orf1ab | C |  |  |  |  |  |  |  |  | T (I) | T (I) |
| 6 | 8344 | orf1ab | C |  |  |  |  |  |  |  | T (D) |  |  |
| 7 | 8782 | orf1ab | C | T (S) | T (S) | T (S) | T (S) |  | T (S) |  |  | T (S) | T (S) |
| 8 | 9962 | orf1ab | C |  |  |  |  |  |  |  |  |  | T (H->Y) |
| 9 | 11430 | orf1ab | A |  |  |  |  |  |  |  |  | G (Y->C) | G (Y->C) |
| 10 | 11916 | orf1ab | C |  |  |  |  |  |  | T (S->L) |  |  |  |
| 11 | 15324 | orf1ab | C |  |  |  |  | T (N) |  |  |  |  |  |
| 12 | 21676 | S | C |  |  |  |  |  |  |  |  | T (Y) | T (Y) |
| 13 | 22081 | S | G |  |  |  |  |  |  | A (Q) |  |  |  |
| 14 | 25672 | ORF3a | C |  |  |  |  |  |  | A (L->I) |  |  |  |
| 15 | 28000 | ORF8 | C |  |  |  |  |  |  | T (P->L) |  |  |  |
| 16 | 28144 | ORF8 | T | C (L->S) | C (L->S) | C (L->S) | C (L->S) |  | C (L->S) |  |  | C (L->S) | C (L->S) |
| 17 | 29095 | N | C |  |  |  |  |  | T (F) |  |  |  |  |
| 18 | 29303 | N | C |  |  |  |  | T (P->S) |  |  |  |  |  |
| 19 | 29625 | ORF10 | C |  |  |  |  |  |  |  |  | T (S->F) | T (S->F) |

L Type (B Lineage)
S Type (A Lineage)
Non- Synonymous
Synonymous
SNP analysis was based on NC\_045512.2 (Wuhan-Hu-1) as the reference genome (Wu et al., 2020). Variants are denoted as nucleotides versus the reference
base. Amino acid changes listed in parentheses with synonymous mutations listed as a single residue. The non-synonymous mutations are bolded.

### SARS-CoV-2 Genotype and Phylogenetic Characteristics

Previous reports demonstrate that SARS-CoV-2 has two genotypes known as L type and S type in the early phase of the outbreak (Tang et al., 2020). The majority of the Shaoxing patients in this study have the S type (70%). We decided to compare our ten SARS-CoV-2 genomes to 163 other published SARS-CoV-2 genomes obtained from GISAID (Shu and McCauley, 2017). Although the majority of the SARS-CoV-2 genomes obtained from GISAID are the L type/B lineage (**Figure 3, red**), the 10 Shaoxing SARS-CoV-2 genomes (**Figure 3, green dots**) are distributed throughout the these genomes with more of them classified in the S type/A lineage (**Figure 3, blue**). Only two outliers (Shenzhen_SZTH-001_2020 and South_Korea_KUMC01_2020) were classified as Lineage A but not assigned to either L or S type (not shown in Figure 3). Due to a lack of the original sequencing data of these two outliers, we are not able to investigate further regarding the discrepancy between the L/S type vs. A/B lineage on them, as either a recombination event or a sequencing error is possible.

**Figure 3.**
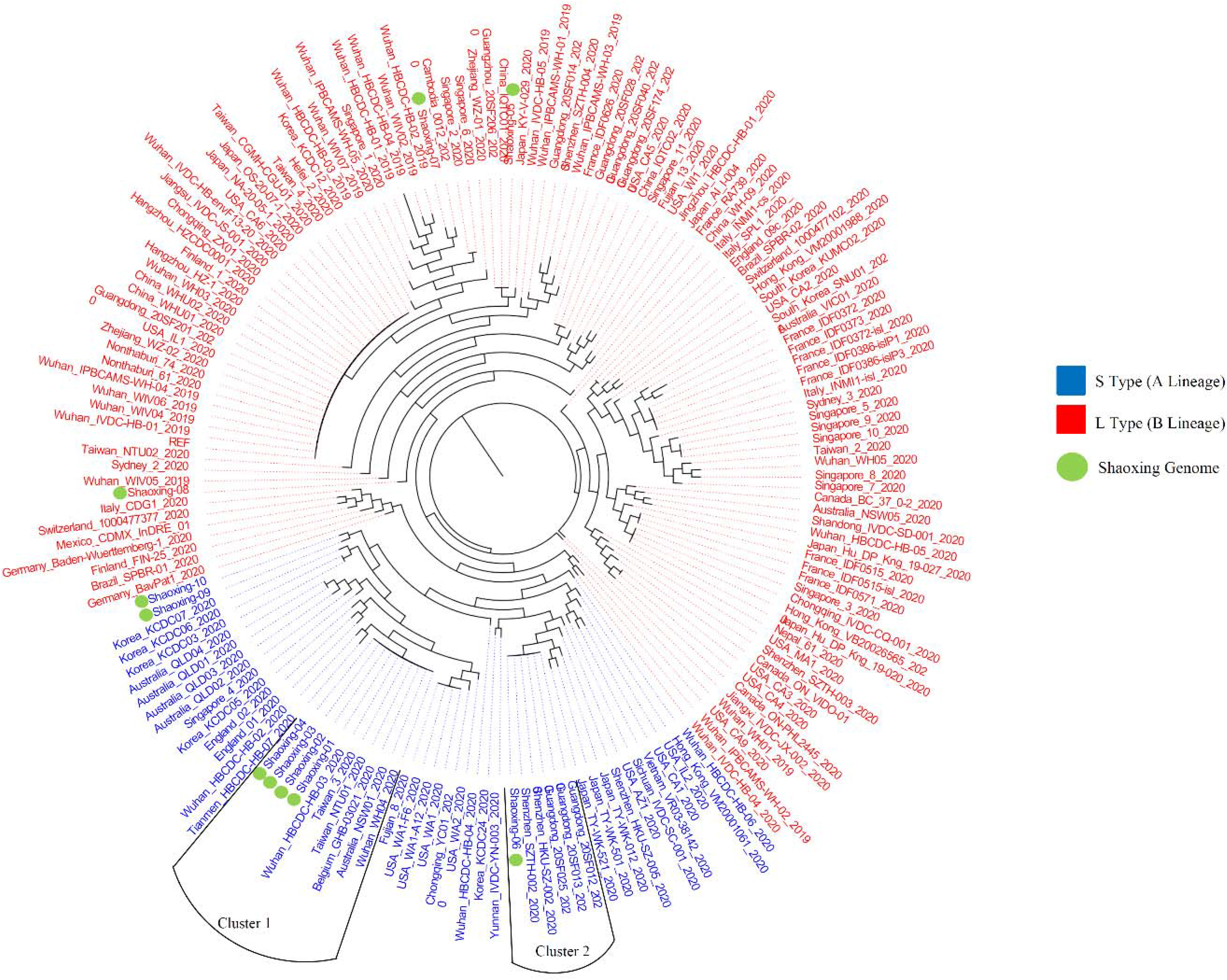
Phylogenetic Comparison of SARS-CoV-2 Genomes. Phylogenetic comparison of 162 published genomes from GISAID (Shu and McCauley, 2017) and the ten Shaoxing genomes (green dots). Genomes are color-coded based on their type/lineage (blue=S type/A lineage, red=L type/B lineage).

Interestingly, four of the Shaoxing SARS-CoV-2 genomes (Shaoxing −1 to −4) were identical to six other GISAID SARS-CoV-2 genomes (**Figure 3, Cluster 1**). These six other genomes were isolated from patients all over the world: two from Wuhan, two from Taiwan, one from Belgium and one from Australia (Figure 3, Cluster 1). Shaoxing-6 is identical to five other genomes isolated in Shenzhen, Guangdong Province in Southern China (**Figure 3, Cluster 2**). Notably, in all ten Shaoxing patients, we found no virus with D614G Spike gene mutation, which was shown to start spreading in Europe in early February, and rapidly become the dominant form in the rest of the world out of China (Korber et al., 2020).

## DISCUSSION

In this study, we sequenced the SARS-CoV-2 genome from ten patient samples from Shaoxing, Zhejiang, China. Using metagenomic sequencing, we were able to obtain above 99% coverage and an average depth of 296X for all 10 SARS-CoV-2 genomes. Although not statistically significant, there does appear to be a clear negative correlation between the C_t_ values of both gene targets and the log count of SARS-CoV-2 RNA sequence reads acquired by metagenomics sequencing. This suggests that the log value of RNA sequence reads by metagenomics sequencing may be used as a semi-quantitative measurement for SARS-CoV-2 viral loads.

The rapid spread of this virus is highlighted by the fact that four SARS-CoV-2 genomes from Shaoxing individuals were identical to six other SARS-CoV-2 genomes from patients all over the world. Our data support recent publications that the virus had spread rapidly around the world especially in Europe before the United States (Deng et al., 2020; Gonzalez-Reiche et al., 2020; Schuchat, 2020).

Overall, we did not see a large number of SNPs in these SARS-CoV-2 genomes. The greatest number of SNPs seen was 9 and these two SARS-CoV-2 genomes were from individuals with no travel history to Hubei province (**Table 3, Shaoxing-9 and 10**). Instead, Shaoxing-9 and 10 had contact with a confirmed case from Ningbo, another city outside of Hubei. We can use these data to infer that the virus accumulated more mutations when it was spread to another city outside of Hubei first before coming to Shaoxing, compared to the virus from people traveled to Shaoxing directly from Hubei.

We combined epidemiologic data with the SNP analysis to estimate the mutation rate of the SARS-CoV-2 from these ten patients. We saw an average mutation rate of 1.37×10^-3^ nucleotide substitution per site per year for SARS-CoV-2, which is consistent with other reports on the mutation rate of SARS-CoV-2(Li et al., 2020;Wang et al., 2020b) and SARS-CoV-1 with a reported mutation rate of 0.80-2.38 × 10^-3^ nucleotide substitution per site per year (Zhao et al., 2004). These data demonstrate that SARS-CoV-2 is similar in the mutation rate as other coronaviruses.

The major limitation of this study is that we only had 10 samples analyzed due to the requirement of sufficient SARS-CoV-2 RNA from a metagenomic sample. However, with the development of a SARS-CoV-2 probe enrichment or multiplex PCR protocols, this type of viral sequencing analysis may be applied to samples with lower viral loads, thereby enabling more complete molecular epidemiological surveillance. In addition, the C_t_ value cut-off of 28 established in this study may not be directly applicable to other real-time PCR assays due to the technical differences.

In summary, we demonstrated that a full viral genomic analysis is feasible via metagenomics sequencing directly on nasopharyngeal samples, which allows retrospective molecular surveillance on SARS-CoV-2 to understand the dynamics of the outbreak in the early phase. The identical virus found in patients in Shaoxing, a mid-sized city outside of Hubei, China, and patients in Europe and Australia was striking. Our analysis added to the growing body of evidence that SARS-CoV-2 spread extremely quickly around the globe as early as January. Although only ten patients were included in this study, we found both lineages (A & B) /types (L & S) of viruses with numerous mutations (both synonymous and non-synonymous) across the entire viral genome. Our study contributed to the understanding of the SARS-CoV-2 evolution in the early phase of the COVID-19 pandemic.

## Data Availability

The datasets and code used for the current study are available from the corresponding author on reasonable request. Example code used for the analysis is available on GitHub.

https://github.com/epimath/cm-dag

## ACKNOWLEDGEMENTS

We would like to thank Yong-Zhen Zhang (Fudan University) and Eddie Holmes (University of Sydney) for sharing the sequence of the first SARS-CoV-2 isolate in a very timely manner. We would also like to thank Fanchao Meng, Bin Hu, Haihao Shou and Yuanyuan Cai from Shaoxing IngeniGen XMK Biotechnologies, Inc. for their technical assistance. This manuscript has been released as a pre-print at MedRxiv: https://doi.org/10.1101/2020.04.16.20058560. (Chen et al., 2020)

